# Forecasting the peak of novel coronavirus disease in Egypt using current confirmed cases and deaths

**DOI:** 10.1101/2020.05.31.20118182

**Authors:** Wagida A. Anwar, Amany Mokhtar

## Abstract

The first case of coronavirus disease 2019 (COVID-19) in Egypt was reported on 14 February 2020 and the number of reported cases has increased daily. The purpose of this study is to describe the current situation of Covid-19 in Egypt and to predict the expected timing of the peak of this epidemic using current confirmed cases and deaths. We used one of the online tools; the Epidemic Calculator that utilizes, the well-known SEIR compartmental model. We utilized the daily reports published by the Egyptian Ministry of Health & Population from 14 February to 11 May 2020. Given the highest calculated case fatality rate (7.7%), the number of hospitalized individuals is expected to peak in the middle of June with a peak of hospitalized cases of 20,126 individuals and total expected deaths 12,303. We recommend strengthening of the Egyptian preventive and control measures so as to decrease the CFR and the number of cases to the least possible as we approach the epidemic peak. It is most important that appropriate quarantine measures are retained., the quarantine measures should not be relaxed before the end of June, 2020.

## Introduction

A pneumonia of unknown cause detected in Wuhan, China was first reported to the WHO Country Office in China in December 2019. On 12 January 2020, the World Health Organization (WHO) confirmed that a novel coronavirus was the cause of this respiratory illness. The outbreak of the disease is ongoing worldwide and the World Health Organization named it coronavirus disease 2019 (COVID-19) on 11 February[1].

COVID-19 is affecting 213 countries and territories around the world. According to WHO, more than 2,900.000 people have been sickened and more than 200,000 people have died of the coronavirus since start of January [2].

The first case of COVID-19 in Egypt was confirmed on 14 February 2020. As of 28 April 2020 there have been 5,042 confirmed cases, 1304 recovered and 359 deaths [3].

The virus is very rapidly spreading. It spread primarily through droplets of saliva or discharge from the nose when infected person coughs or sneezes. Studies to date suggest that the virus that causes COVID-19 transmits mainly through contact with respiratory droplets rather than through air. The virus may be present in feces of some cases but spread through this route is not the main feature of the outbreak. However the transmission of the virus and progress of the disease in people of different ages remain an area of emerging research [4].

Limited testing and other challenges in the attribution of the cause of death means that the number of confirmed deaths may not be accurate. Now there is an evidence that people who do not show sever symptoms can spread it silently also there is slow rollout of diagnostic tests. So we don’t have a precise case count or know where the virus might spread [5].

Infectious disease pandemics in recent years have increased interest in real-time forecasting and long-term prediction of epidemic curves and disease transmission parameters [6]. It is an important indication for the severity of an epidemic as a function of time. The cumulative cases during the initial growth phase form an approximately linear relationship with time in log-linear scale. Thus, in linear scale, the number of deaths increases exponentially with time. [7].

A good estimate of the epidemic curve during an outbreak would be valuable to health care officials. Based on this estimate, they can plan for sufficient resources and supplies to handle disease treatment on a timely basis. When we say we are estimating the epidemic curve during an outbreak, we mean that, on a given day of the outbreak, we are estimating the daily number of new outbreak cases for days that have occurred so far and we are predicting those daily values for days that will occur in the future [8].

Disease progression modeling involves the utilization of mathematical functions as quantitative descriptors of the time course of disease status. Disease models may incorporate biomarkers of disease severity and/or clinical outcomes to characterize the natural progression of disease[9], however, for an emergent disease, such as Covid-19, with no past epidemiological data to guide models, modelers struggle to make predictions of the course of the pandemic. Policy decisions depend on such predictions but they vary widely [10].

The ability to predict the turning points and the end of the epidemic is of crucial importance for fighting the epidemic and planning for a return to normality. The accuracy of the prediction of the peaks of the epidemic is validated using data in different regions in China showing the effects of different levels of quarantine. The validated tool can be applied to other countries where Covid-19 has spread, and generally to future epidemics [10].

This study was designed to describe the current situation of Covid-19 in Egypt and to give a prediction of the expected timing of the peak of this epidemic in Egypt using one of the online tools; the Epidemic Calculator that utilizes, the well-known SEIR compartmental model susceptible, exposed, infective and removed populations at time t.

## Material and Methods

### 1 Situational analysis of COVID –19 pandemic in Egypt

To describe the current situation of the pandemic in Egypt, we utilized the daily reports published by the Egyptian Ministry of Health & Population (MoHP). New cases, new deaths, total deaths, total recovered, and total cases were recorded and we calculated the case fatality rate (CFR) by dividing the total deaths by the total number of confirmed cases.

We draw the epidemic curve for the new cases, total cases, total recovered and total deaths using Microsoft excel. The logarithmic scale was used that could give a more detailed description of the situation. Recovery rate was calculated by dividing the reported total recovered cases by the total number of confirmed cases.

### 2 Prediction of the epidemic peak for COVID-19 in Egypt

The classical susceptible exposed infectious recovered model (SEIR) is the most widely adopted one for characterizing the epidemic of COVID-19 in both China and other countries [11]. Based on SEIR model, one can also assess the effectiveness of various measures since the outbreak, which seems to be a difficult task for general statistics methods [12].

Prediction of the epidemic peak for COVID-19 in Egypt can be done by using the real-time data from 14 February to date. Taking into account the uncertainty due to the incomplete identification of infective population, we can apply the well-known SEIR compartmental model for the prediction based on daily observations.

We used the Epidemic Calculator developed by Gabe, 2020.This calculator implements the **SEIR** model, the idealized model of spread still used in frontlines of research [13][14].The dynamics of this model are characterized by a set of four ordinary differential equations that correspond to the stages of the disease’s progression[15].

According to the epidemic calculator, the following parameters were used:

1. **Transmission Dynamics**

a. ***Population Inputs:***

i. Size of population: 100,000000
ii. Number of initial infections = 1
b. ***Basic Reproduction Number R_0_*:** Measure of contagiousness: the number of secondary infections each infected individual produces. The R_0_ used in this equation was set at 2.3 ([16]
c. ***Transmission Times***

i. Length of incubation period, *T*_inc_; we set it at (5.2 days) [16]
ii. Duration patient is infectious, *T*_inf_. ; was estimated to be 4 days [16]
2. **Clinical Dynamics**

a. **Mortality Statistics**

i. Case fatality rate (CFR); we used different values for the CFR which were calculated using the published data by the Egyptian Ministry of Health and population from March 08,2020 till April 28,2020; mean case fatality rate in days for the entire duration (5.6 % ± 2.1 %SD) with 95% CI (5.0- 6.3)
ii. Time from end of incubation to death.;33 days [16]
b. **Recovery Times**[16]

i. Length of hospital stay: 28.6 days
ii. Recovery time for mild cases: 11.1 days
c. **Care statistics**

a. Hospitalization rate; was set at 5 % [17]
b. Time to hospitalization: 5 days [16]

## Model Details

The clinical dynamics in this model are an elaboration on SEIR that simulates the disease’s progression at a higher resolution, subdividing I&R into mild (patients who recover without the need for hospitalization), moderate (patients who require hospitalization but survive) and fatal (patients who require hospitalization and do not survive). Each of these variables follows its own trajectory to the final outcome, and the sum of these compartments add up to the values predicted by SEIR. .We assume, for simplicity, that all fatalities come from hospitals, and that all fatal cases are admitted to hospitals immediately after the infectious period [15].

## Results

The first case of coronavirus disease 2019 (COVID-19) in Egypt was reported on 14 February 2020 and the number of reported cases has increased day by day. There was an obvious increase in the number of reported cases, total deaths, and also increase in the number of recovered cases Figure (1). However, looking at the epidemic curves (the logarithmic graphs of COVID-19) we can observe the slow rate of growth even though the overall numbers are still increasing (Figures 2&3). As of April 28, the recovery rate was 25.9 and the CFR was 7.1% (Figures 4&5)

**Figure 1:**
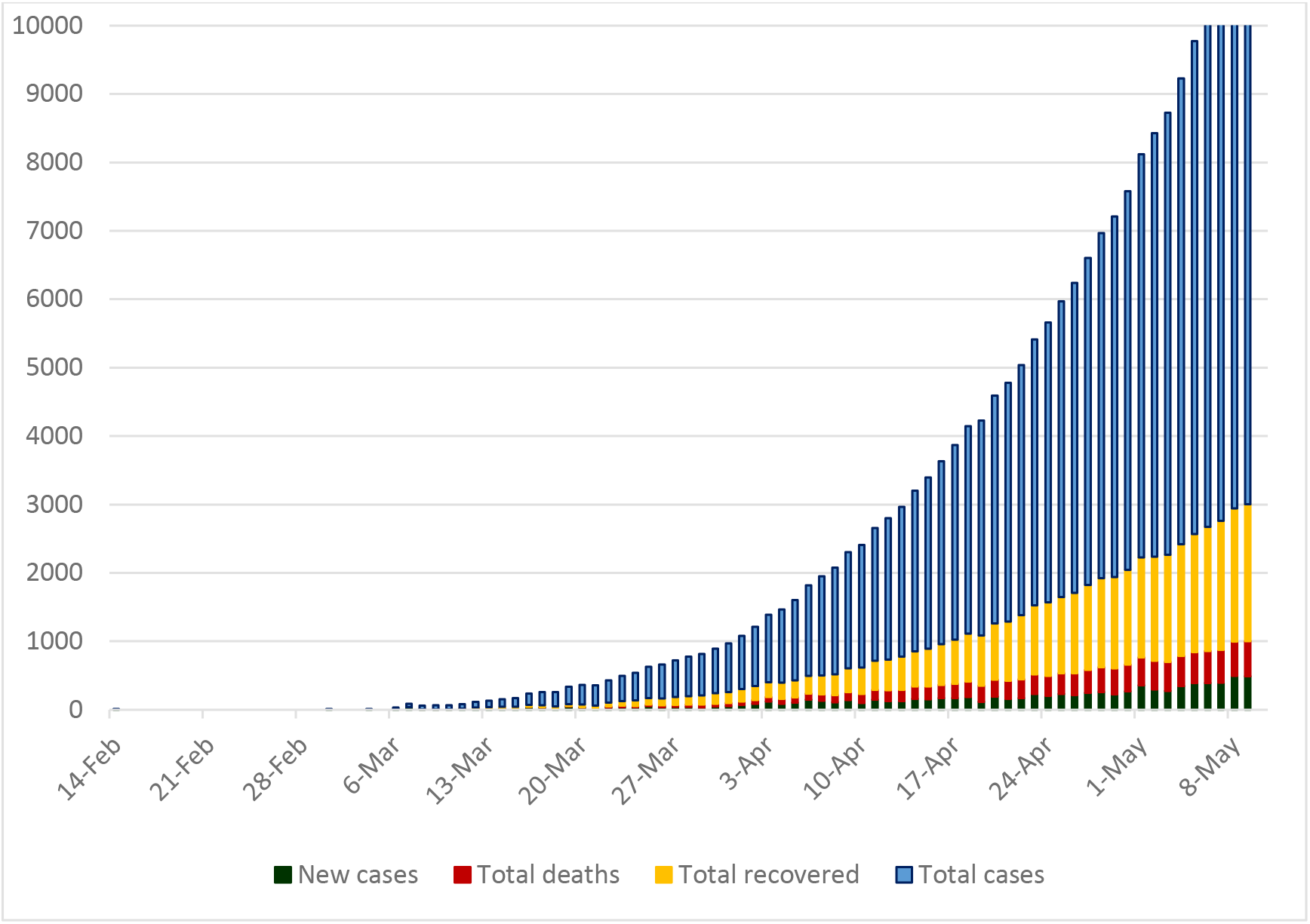
Number of COVID-19; new cases, total deaths, total recovered and total cases in Egypt

**Figure 2.**
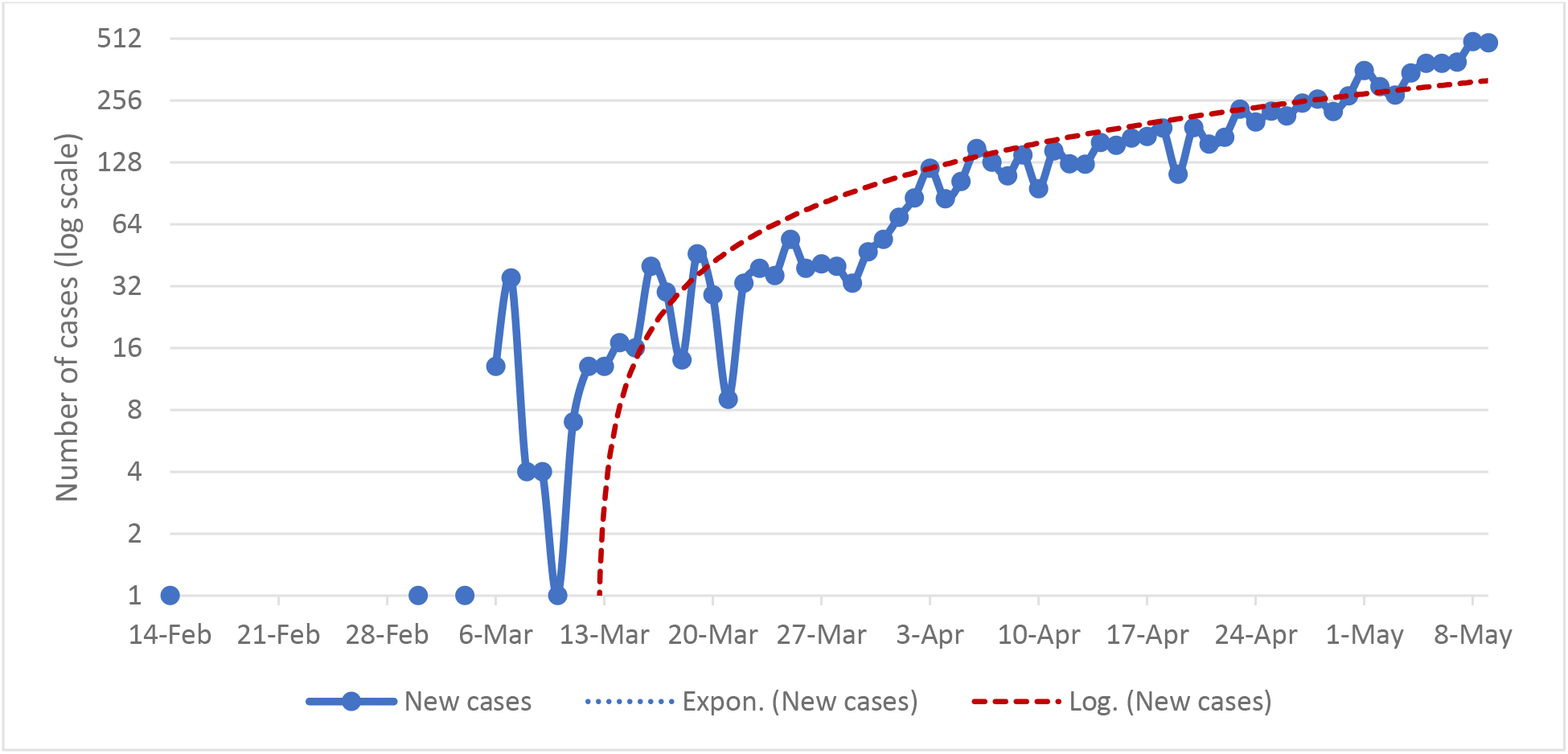
Epidemic curve of new COVID-19, cases through 9, May2020

**Figure 3.**
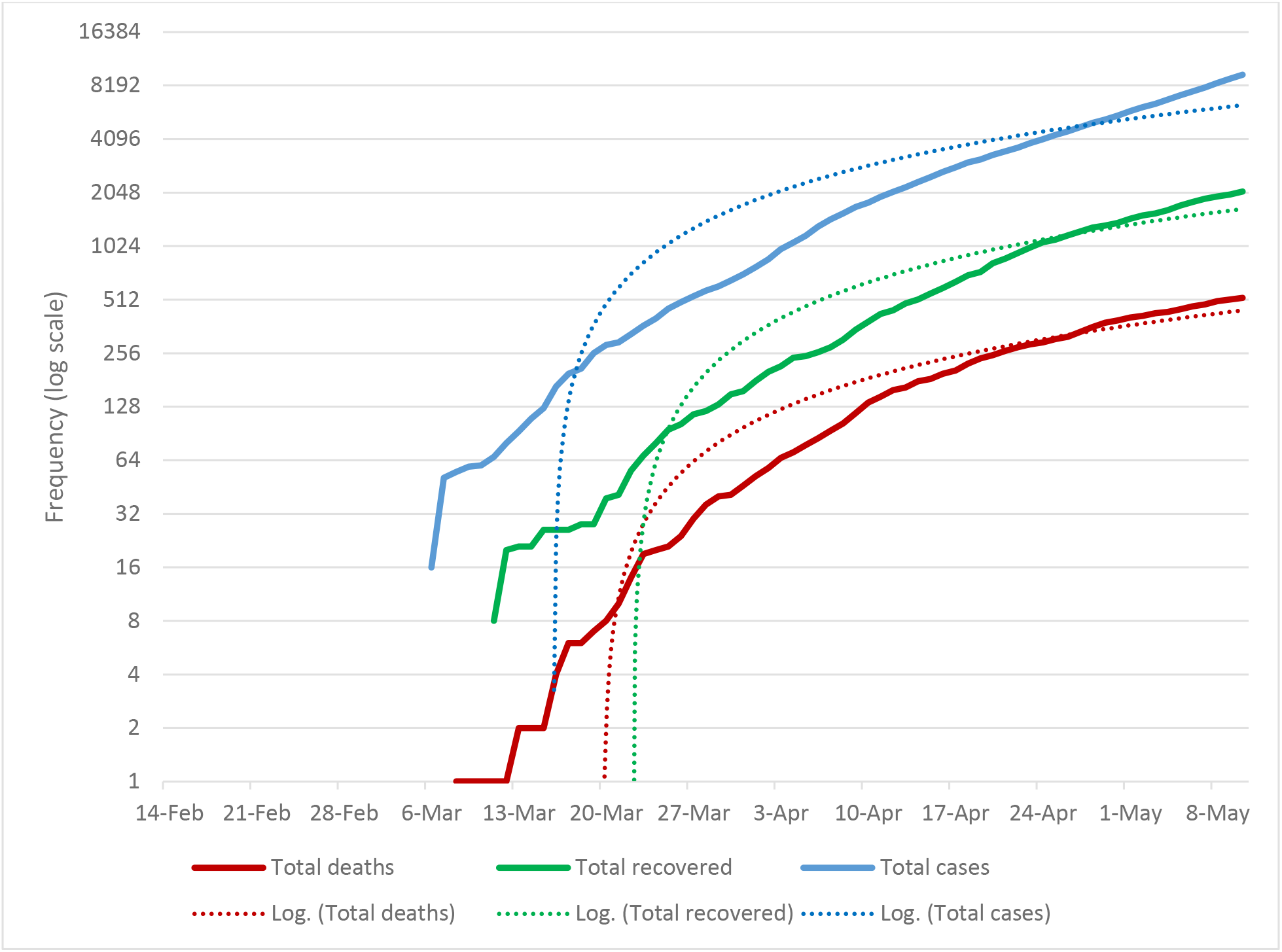
Epidemic curve of COVID-19; total cases, total recovered and total deaths

**Figure 4.**
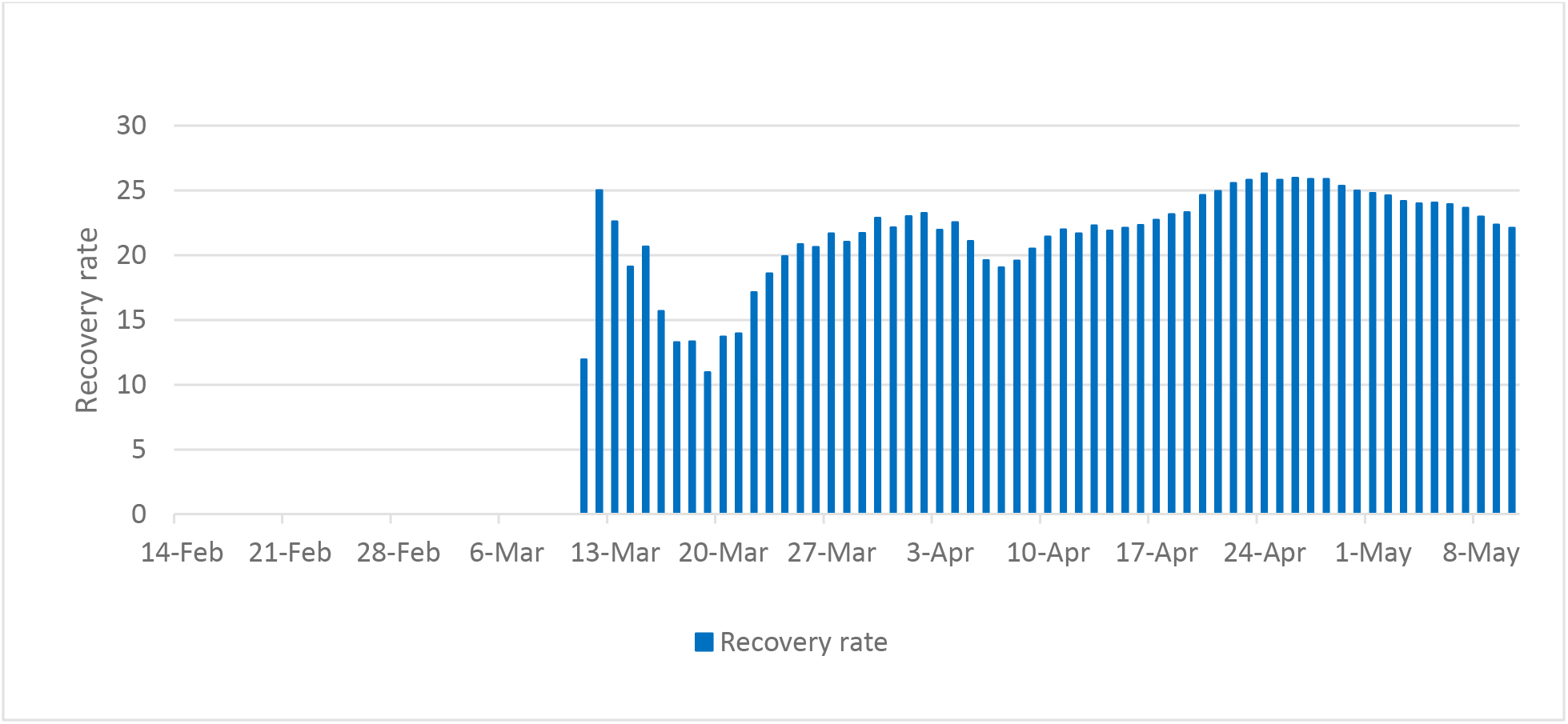
Recovery rate of COVID-19 cases in Egypt

**Figure 5.**
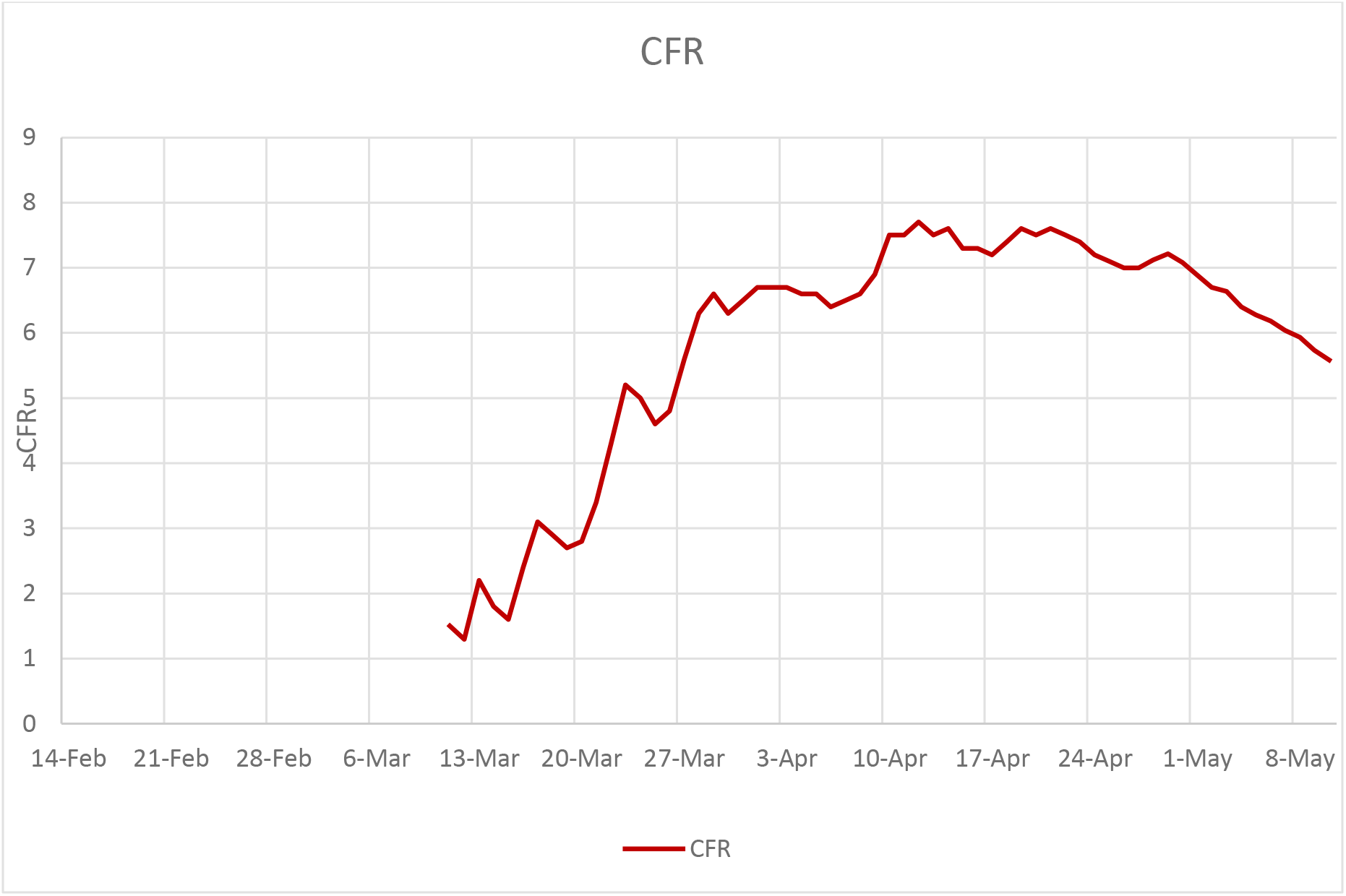
Case fatality rate of COVID-19 pandemic in Egypt

Simulations results ran for the previously described parameter combinations and for the calculated case fatality rates are summarized in Table 1. Whilst there is substantial uncertainty in the total numbers of cases, results under most settings suggest that hundreds to thousands of cases are likely. Higher reproduction numbers (*R*) and lower CFR give the largest estimates. The scenario leading to fewest cases (*R* = 1.5, CFR = 10%), which may be reasonable for deaths in elderly patients (results not shown), still suggest that a few thousands of contacts (assuming a few 10s of contacts per case) would likely have to be followed in order to contain the epidemic.

**Table 1:** Summary table of the predicted peak of hospitalization at different CFR

| CFR | Peak hospitalization number | predicted date of peak | Total expected deaths |
| --- | --- | --- | --- |
| 5.6 | 16,789 | 12-Jun-20 | 8,947 |
| 6.6 | 18,378 | 14-Jun-20 | 10,545 |
| 6.9 | 18,855 | 14-Jun-20 | 11,024 |
| 7.1 | 19,173 | 14-Jun-20 | 11,344 |
| 7.5 | 19,808 | 14-Jun-20 | 11,983 |
| 7.7 | 20,126 | 16-Jun-20 | 12,303 |

Using the epidemic calculator at different CFR values resulted in predicting nearly the same date of hospitalization peak (12–16 June, 2020), moreover increasing the CFR resulted in predicting higher numbers of cases at the peak (Figure 6)

**Figure 6.**
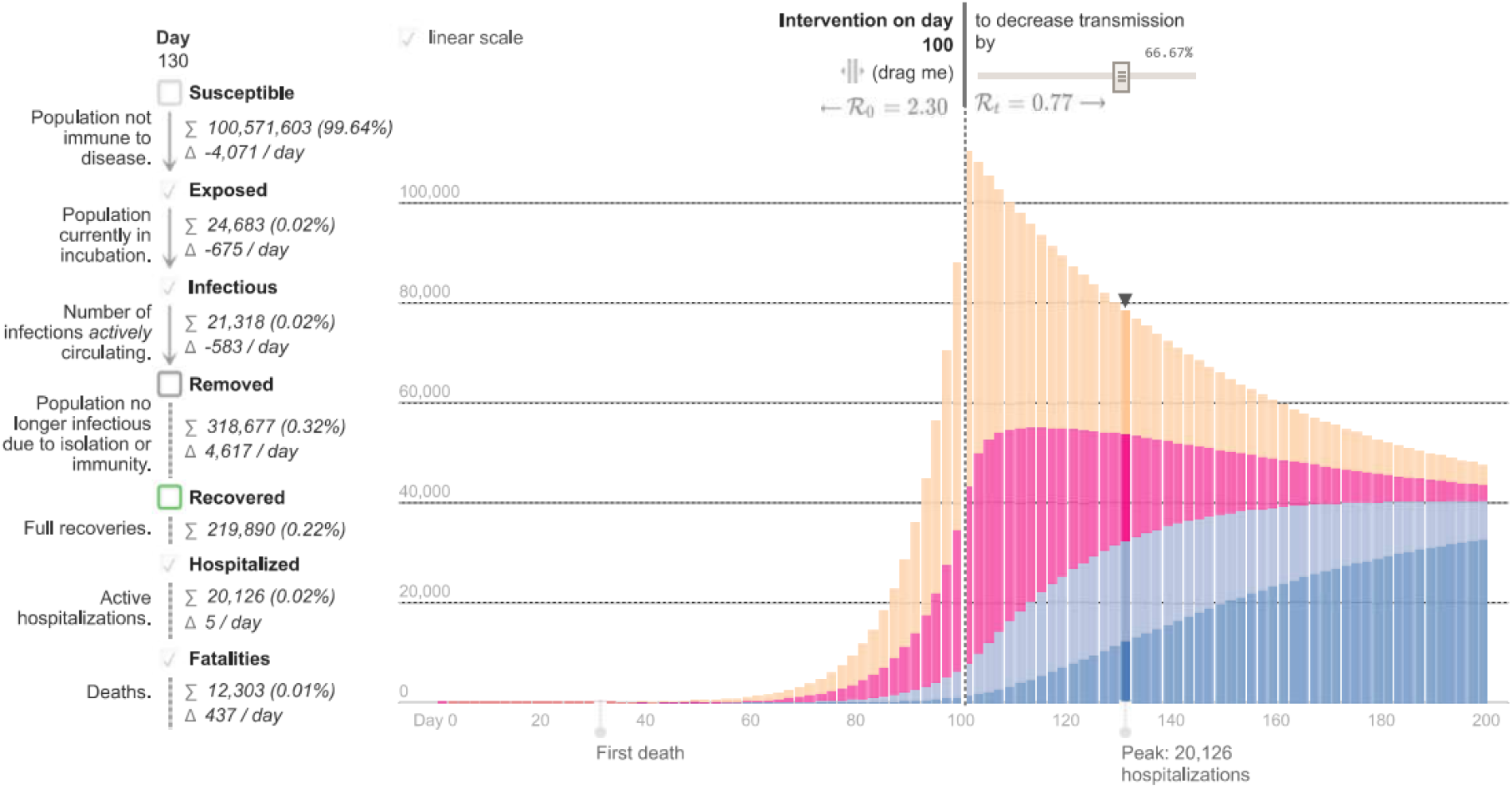
Predicted peak of hospitalization at case fatality rate 7.7 %

## Discussion

The number of people who have ever tested positive for coronavirus in a given country, regardless of whether they have recovered, can illustrate how the pandemic is expanding. An upward bend in a curve can indicate either a time of explosive growth of coronavirus cases in a given country or a change in how cases are defined or counted. Comparing across countries can also show where the pandemic is growing most rapidly at any point in time [18].Moreover, daily new cases data provides an early indication as to whether a country’s epidemic curve is flattening and if containment measures are working. They are also a good estimate of the current impact on the health-care system. According to our data the epidemic curve in Egypt is slowly ascending as of April, 28. Representation of the epidemiological curve depend on many factors including, the availability and access to the COVID-19 testing; not all countries have the capacity or the will to test for cases extensively or conduct effective contact tracing to find cases in the community, and it can be difficult to verify each country’s data. The population response to the public health interventions, the magnitude and the strength of the mitigation actions, the response of the immune system, the mutations of the virus, and the seasonality of the spread of the infection should also considered [19].

In this study, we propose the SEIR compartmental model to predict the epidemic peak of COVID-19 using one of the online tools; the Epidemic Calculator. Given the highest calculated case fatality rate, the number of hospitalized individuals is expected to peak on 16^th^ June with a peak of hospitalized cases of 20,126 individuals and total expected deaths 12,303. A study conducted by El-Ghitany, 2020 expected the active cases to reach more than 20,000 by late May then starts to decline [20] while, another study conducted by the tachyhealth team expected the peak to be between 6–18 June 2020.

Based on our study we recommend strengthening of the Egyptian preventive and control measures so as to decrease the CFR and the number of cases to the least possible as we approach the epidemic peak. It is most important that appropriate quarantine measures are retained., the quarantine measures should not be relaxed before the end of June, 2020.

Modeling and estimation of the epidemic trend and the effect of intervention can be enhanced by improvement of data quality that requires collaboration between Ministry of Health and population and other sectors to get the required data

### limitations of modeling the situation of COVID-19

- Much is still unknown or uncertain about the virus

- The lag time in research and publication of journals means understanding of the virus is moving faster than refereed research
- The estimated model is built using standard epidemiological modeling techniques, but given the relatively early stage of our understanding of this virus, it is possible that the virus does not behave in a way that makes such techniques applicable
- The transmission of the virus and progress of the disease in people of different ages remain an area of emerging research. This version of the model does not yet and other sectors to get the required data incorporate an age stratification or other features that correct for differing demographics between geographies.
- There are very significant differences in access of testing and rates of testing and /or the timelines and reliability of the reporting of infections across different geographies

- The availability and quality of data will have a material impact on the quality and reliability of outputs
- Government policy interventions have a significant lag-time

- Given the time between infection, incubation, development of symptoms, access to testing and results, the impact of a particular government policy intervention taken today is unlikely to change the shape of the curve for at least one week, and possibly materially longer.

## Data Availability

All data referred to in the manuscript are available in the supplementary file

## Notes

### Competing Interest Statement

The authors have declared no competing interest.

### Funding Statement

no external funding was received

### Author Declarations

We utilized the daily reports published by the Egyptian Ministry of Health & Population online and it is available for every one, so we are not required to get approval from our university to conduct our research

## References

1. Kuniya T. Prediction of the Epidemic Peak of Coronavirus Disease in Japan, 2020. J Clin Med. 2020 Mar;9(3).

2. WHO. Coronavirus disease (COVID-19) outbreak situation [Internet]. 2020 [cited 2020 Apr 26]. Available from: https://www.who.int/emergencies/diseases/novel-coronavirus-2019

3. Ministry of health and population. COVID 19 [Internet]. 2020 [cited 2020 Apr 26]. Available from: https://www.care.gov.eg/EgyptCare/News/Browse.aspx?cid=1

4. Verity R, Okell LC, Dorigatti I, Winskill P, Whittaker C, Imai N, et al. Estimates of the severity of coronavirus disease 2019: a model-based analysis. Lancet Infect Dis. 2020 Mar;

5. CDC. COVID-19 News Search Alert [Internet]. 2020 [cited 2020 Mar 28]. Available from: https://www.cdc.gov/library/researchguides/2019novelcoronavirus/newssearchalert.html

6. Nsoesie EO, Beckman R, Marathe M, Lewis B. Prediction of an Epidemic Curve: A Supervised Classification Approach. Stat Commun Infect Dis. 2011;

7. Ma J. Estimating epidemic exponential growth rate and basic reproduction number. Infectious Disease Modelling. 2020.

8. Jiang X, Wallstrom G, Cooper GF, Wagner MM. Bayesian prediction of an epidemic curve. J Biomed Inform. 2009;

9. Cook SF, Bies RR. Disease Progression Modeling: Key Concepts and Recent Developments. Curr Pharmacol reports [Internet]. 2016/08/15. 2016 Oct;2(5):221–30. Available from: https://pubmed.ncbi.nlm.nih.gov/28936389

10. Huang NE, Qiao F, Tung K-K. A data-driven tool for tracking and predicting the course of COVID-19 epidemic as it evolves. medRxiv. 2020;

11. Read JM, Bridgen JRRE, Cummings DATA, Ho A, Jewell CP. Novel coronavirus 2019-nCoV: early estimation of epidemiological parameters and epidemic predictions. medRxiv. 2020;

12. Peng L, Yang W, Zhang D, Zhuge C, Hong L. Epidemic analysis of COVID-19 in China by dynamical modeling. arXiv Prepr arXiv200206563. 2020;

13. Wu JT, Leung K, Leung GM. Nowcasting and forecasting the potential domestic and international spread of the 2019-nCoV outbreak originating in Wuhan, China: a modelling study. Lancet. 2020;

14. Kucharski AJ, Russell TW, Diamond C, Liu Y, Edmunds J, Funk S, et al. Early dynamics of transmission and control of COVID-19: a mathematical modelling study. Lancet Infect Dis. 2020;

15. Gabgoh. Epidemic Calculator [Internet]. 2020 [cited 2020 Apr 14]. Available from: https://gabgoh.github.io/COVID/index.html

16. Github. COVID-19/parameter_estimates/2019_novel_coronavirus [Internet]. 2020 [cited 2020 Mar 25]. Available from: https://github.com/midas-network/COVID-19/blob/master/parameter_estimates/2019_novel_coronavirus/README.md

17. Wu Z, McGoogan JM. Characteristics of and Important Lessons From the Coronavirus Disease 2019 (COVID-19) Outbreak in China: Summary of a Report of 72 314 Cases From the Chinese Center for Disease Control and Prevention. JAMA [Internet]. 2020 Apr 7;323(13):1239–42. Available from: https://doi.org/10.1001/jama.2020.2648

18. Johns Hopkins University. MAPS & TRENDS [Internet]. 2020 [cited 2020 Apr 25]. Available from: https://coronavirus.jhu.edu/data/cumulative-cases

19. tachyhealth. Predicting the peak of the epidemic curve of COVID-19 in selected Arab countries [Internet]. 2020 [cited 2020 Apr 23]. Available from: https://www.tachyhealth.com/blog/predicting-the-peak-of-the-epidemic-curve-of-covid-19-in-selected-arab-countries

20. El-Ghitany E. A Short-Term Forecast Scenario for COVID-19 Epidemic and Allocated Hospital Readiness in Egypt. Preprints. 2020

